# Effectiveness of Traditional Chinese Exercises for Limb Function Rehabilitation after Stroke: A Systematic Review and Meta-analysis

**DOI:** 10.1101/2025.03.24.25324568

**Authors:** Wang Guo, Hong-yu Li, Song-qi Li, Xuan-ting Chen, Zhi-hao Wang, Qiang Tang

## Abstract

**Background:** As a form of exercise, Traditional Chinese Exercises (TCEs) have shown unique advantages in the stroke population. TCEs have been indicated by existing research to potentially improve limb function and activities of daily living (ADL) in stroke survivors. Still, a unified analysis of these results has been lacking. This systematic review seeks to assess the therapeutic impact of six specific types of TCEs— Daoyin, Tai Chi, Yijinjing, Baduanjin, Liuzijue, and Wuqinxi—on limb motor function, balance function, physical function, muscle tone and and ADL in stroke patients.

**Methods:** Adhering to the PRISMA Guidelines (CRD42024605971) and the P.I.C.O.S. Standard, RCT studies were extracted from eight databases, comprising four Chinese and four English ones. Risk of bias and quality of evidence were assessed using RoB2 and GRADE tools. The quality of the randomized controlled trials (RCTs) was evaluated, and a meta-analysis was conducted using RevMan 5.4.

**Results:** There were 28 RCTs analyzed by this systematic review. The meta-analysis indicated that TCEs yielded significant therapeutic benefits in improving motor function (SMD = 1.05, 95% CI: [0.70 to 1.40], *P* < 0.001), balance function (SMD = 0.81, 95% CI: [0.43 to 1.18], *P* < 0.001), muscle tone (MD = -0.46, 95% CI: [-0.69 to -0.24], *P* < 0.001), and ADL (MD = 5.53, 95% CI: [4.37 to 6.69], *P* < 0.001). Subgroup analysis showed that Taiji, Baduanjin and Yijinjing had significant rehabilitation effects in these aspects during 2-12 weeks of treatment (all *P* < 0.05).

**Conclusions:** TCEs improved stroke patients’ limb motor and balance function. They also reduced muscle tension, enhanced daily activity ability, and demonstrated high safety. Tai Chi, Yijinjing, and Baduanjin showed unique advantages of rehabilitation treatment in these aspects. More high-quality, multicenter RCTs are needed to further confirm these results.

## Introduction

Stroke represents a severe cerebrovascular disorder that poses a significant threat to human health. In developing countries, the incidence of stroke is expected to rise sharply over the next several decades, largely due to demographic changes, especially the aging population[1]. Globally, stroke ranks as the third most common cause of both disability and death[2, 3]. Research indicates that by 2030, 3.4 million American adults will experience a stroke, with stroke-related mortality rising by approximately 50%. This is expected to result in an additional 64,000 stroke deaths annually compared to 2012[4]. Beyond the stark mortality statistics, the disabling effects of stroke are even more profound. According to the American Heart Association, nearly 75% of stroke survivors exhibit mild to moderate disability, while 15% to 30% suffer from severe disability[5]. Even after undergoing rehabilitation, 65% of stroke patients fail to fully regain motor function[6]. Undoubtedly, the societal and familial burdens imposed by stroke are immense.

Traditional Chinese Exercises (TCEs), originating from ancient China, are a form of low-to-moderate intensity aerobic exercise. These practices, which include Daoyin, Tai Chi, Yijinjing, Baduanjin, Liuzijue, Wuqinxi, and Qi Gong[7], not only reflect the profound cultural heritage of China but also encapsulate the foundational principles of Traditional Chinese Medicine[8]. TCEs are designed to be low in intensity, focusing on breathing regulation and meditation to promote relaxation[9]. Unlike high-intensity physical exercise, TCEs serve as a therapeutic method for functional compensation and rehabilitation, enabling the body to adapt to and recover from disease while maintaining overall bodily function[10]. TCEs offer numerous advantages as a non-pharmacological and non-invasive therapeutic option. They are easy to practice, highly accessible, and simple to implement, making them particularly suitable for rehabilitation in TCM, especially for cerebrovascular diseases. In stroke recovery, TCEs have shown significant benefits, including improved motor function, enhanced cognitive recovery, increased physical capability, reduced fall risk, and overall better rehabilitation outcomes[11–14]. These characteristics make TCEs a valuable tool in stroke rehabilitation, setting them apart from more traditional, high-intensity physical therapies.

Research has suggested that certain forms of TCEs may have limited effectiveness in post-stroke rehabilitation, especially in improving balance[15–17]. This underscores ongoing debate in the field. Although numerous systematic reviews and meta-analyses have assessed the effects of TCEs on limb function recovery following a stroke[18–20], these studies have generally focused on specific techniques, often overlooking other forms such as Liuzijue and Wuqinxi. This narrow focus limits the ability to identify the most effective treatment approaches. Given these limitations, a more comprehensive evaluation of the therapeutic role of TCEs in post-stroke rehabilitation is needed. The purpose of this study was to evaluate the effects of TCEs and other rehabilitation methods on limb function in stroke patients (including imb motor function, balance function, physical function, physical function). muscle tone and daily living ability) for all available evidence, and to provide reference for clinical practice and follow-up studies.

## Methods

### Search strategy

Following the PRISMA guidelines for systematic reviews and meta-analyses, this study was rigorously conducted and registered with PROSPERO (Registration No: CRD42024605971). To gather relevant articles, a hybrid method combining computer-assisted and manual searches was employed across multiple databases, including Chinese Scientific Journal Database(VIP), China National Knowledge Infrastructure(CNKI), WanFang Data, the Chinese Biomedical Literature Database, Web of Science, EMBase, The Cochrane Library, and PubMed, covering publications from their establishment through March 13, 2025. Key search terms included“Chinese traditional exercise”, “Yijinjing”, “Wuqinxi”, “Baduanjin”, “Liuzijue”, “Daoyin”, “Tai Ji”, “Qigong”, “Stroke”, “Cerebral Hemorrhage”, “Apoplexy”, and “Randomized Controlled Trial” et al. Search strategies were customized for each database. The English search strategy used MeSH subject words to combine with free words to retrieve publications. A detailed search strategy could be found in supplementary material. Took Tai Ji search in the PUBMED database as an example:

#1: “Tai Ji” [MeSH] OR “Tai Chi” [Title/Abstract] OR “Tai Chi Chuan” [Title/Abstract] OR “Taijiquan” [Title/Abstract].

#2: “Stroke” [MeSH] OR “Ischemic stroke” [Title/Abstract] OR “Cerebral Hemorrhage” [Title/Abstract] OR“Apoplexy” [Title/Abstract].

#3: “Randomized Controlled Trial” [Publication Type]. #4: #1 and #2 and #3.

### Research eligibility

Literature was searched and screened independently by two authors, who also performed data extraction, assessed the risk of bias and evaluated the evidence quality. In the event of a disagreement, a third author was consulted to resolve the issue. In this review, the Cochrane Library handbook version 2 (RoB2) was applied for assessing risk of bias[21]. The GRADE evidence grading and recommendation system was utilized to appraise the quality of evidence[22]. The criteria for inclusion were: (1) randomized controlled trials (RCTs). (2) participants diagnosed with stroke via MRI or CT scans. (3) The TCEs specifically included Yijinjing, Baduanjin, Tai Chi, Liuzijue, Wuqinxi, Qigong, and Daoyin. (4) The intervention group received a single form of TCE, with or without additional treatments. (5) The control group was given standard rehabilitation therapies or exercise interventions, which included acupuncture, conventional rehabilitation, routine treatments, or general health advice. The exclusion criteria were: (1) incomplete or erroneous data, or unavailable studies. (2) Duplicate publications. (3) Reviews and other non-original studies. (4) Participants with cognitive impairments, contraindicated conditions, heart failure, kidney failure, or severe illness.

### Data extraction

The data collected from the included studies included various indicators: (1) Author, publication year, country, sample size, gender, age, interventions, and control measures et al. (2) Primary outcome measures, including the Fugl-Meyer Assessment (FMA), the Fugl-Meyer Assessment for Upper Limb (FMA-U), the Fugl-Meyer Assessment for Lower Limb (FMA-L), the Short Physical Performance Battery (SPPB), Berg balance scale (BBS), the Timed Up and Go Test (TUGT), and the 2-minute step test. (3) Secondary outcome measures, which included the Modified Barthel Index (MBI) and the Modified Ashworth Scale (MAS).

### Data analysis

Data analysis was performed with RevMan 5.4 software. When different assessment tools were utilized, the standardized mean difference (SMD) was applied as the effect measure. For studies using the same tool, the mean difference (MD) was employed. All analyses included the calculation of 95% confidence interval (CI) for effect sizes. The chi-squared test and *I²* statistic were used to assess heterogeneity. Heterogeneity was assessed using the chi-squared test and I² statistic. Non-significant heterogeneity was deemed present if *P* > 0.05 and I² < 50%, prompting the use of a fixed-effects model for the meta-analysis. In cases where *P* < 0.05 and *I²* exceeded 50%, indicating significant heterogeneity, a random-effects model was employed. Subgroup or sensitivity analyses were conducted to explore heterogeneity sources and evaluate result reliability. To assess publication bias, a funnel plot was utilized when at least 10 studies were included.

## Results

### Literature search results

Initially, 5,603 articles were retrieved, with 5,089 remaining after duplicates were excluded. Post-review of titles and abstracts, 4,973 articles were omitted because they did not meet the predefined criteria concerning interventions and study types. Following a full-text assessment of the remaining 116 articles, 88 were eliminated: 59 non-RCTs, 9 with unclear diagnoses, 14 with unsuitable interventions, 6 lacking relevant outcome measures. Ultimately, 28 RCTs (21 in Chinese[23–43] and 7 in English[14–17, 44–46]) were included in the analysis after meeting the inclusion criteria. The review process was depicted in **Fig 1**.

**Fig 1.**
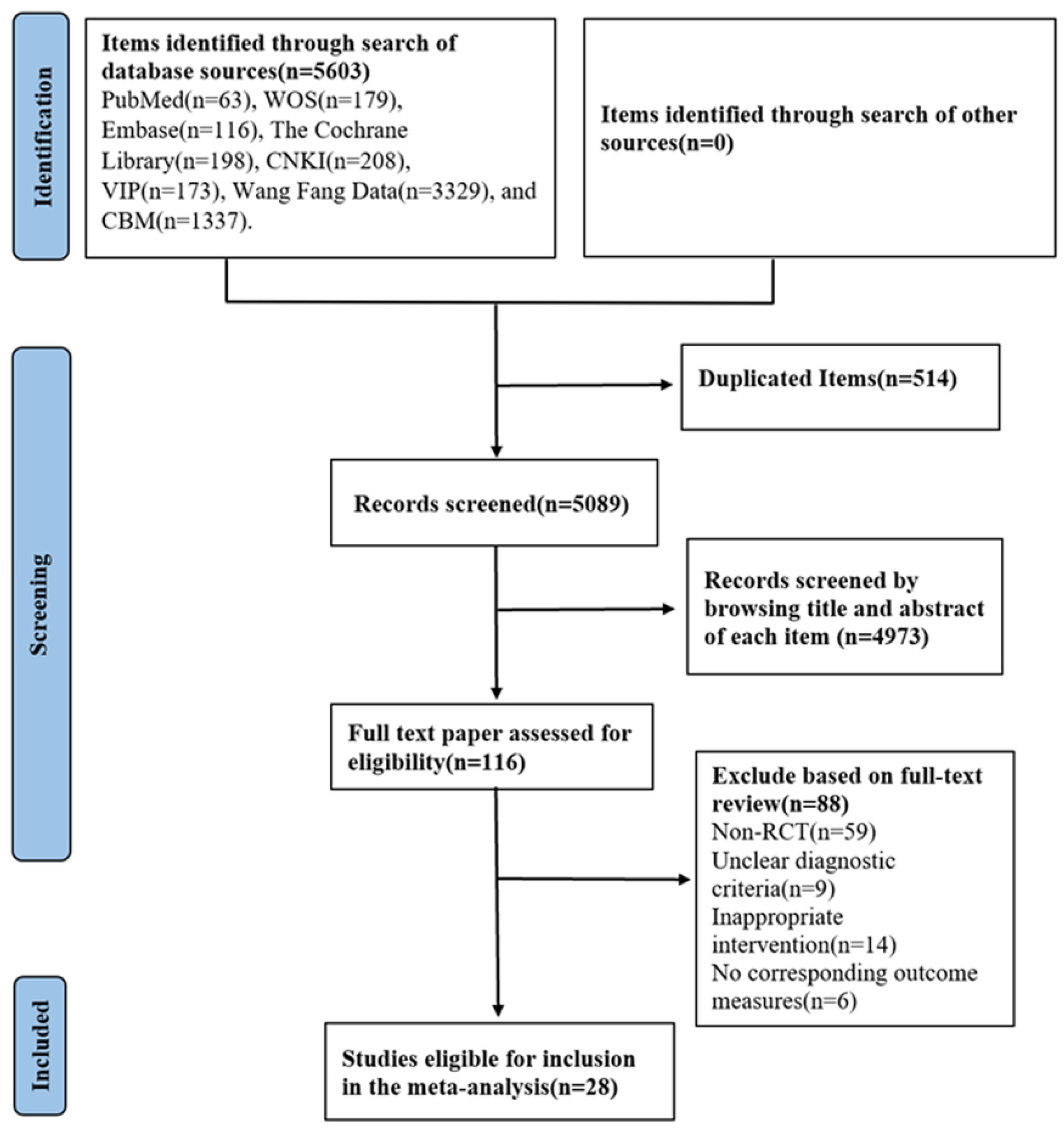
Flow chart of study.

### Study characteristics

**Table 1 and Table 2** showed the basic characteristics of these 28 studies. A total of 1686 stroke patients were included in the study, with 857 participants in the experimental group and 829 in the control group. Among the included studies, 24 originated from China[14–16, 23–43], 2 from the United States[44, 45], and 2 from South Korea[17, 46]. Six types of interventions were employed: 11 studies used Tai Chi[15, 17, 25, 26, 28, 33, 34, 43–46], 4 studies used Baduanjin[23, 35, 36, 42], 7 studies used Yijinjing[24, 30, 33, 37, 39–41], 1 study used Daoyin[16], 3 studies used Liuzijue[14, 27, 31], and 2 study used Wuqinxi[29, 38]. Interventions ranged in duration from 2 to 20 weeks, with 12 weeks being the most frequently applied[15, 17, 24, 30, 31, 33, 36, 37, 39, 43–45]. Most interventions were conducted five times per week. Motor function was primarily assessed using the FMA, FMA-L, and FMA-U scales[16, 23, 25–27, 29, 30, 32–37, 39]. Seventeen studies used balance function tests, such as the BBS or TUGT[15–17, 23–28, 33, 36, 38, 40–43, 46]. Nine studies assessed daily living abilities using the MBI scale[14, 24–27, 31, 32, 39, 40]. Additionally, 2 studies evaluated physical function[44, 45], and 2 others measured muscle tone[26, 32]. While only 6 studies addressed the safety of TCEs[17, 26, 34, 38, 41, 45]. One study reported a case of a patient who dropped out of the trial with dizziness[41].

**Table 1.** The basic characteristics of studies.

| Author | Country | Study design | Experimental group |  |  |  | Control group |  |  |  |
| --- | --- | --- | --- | --- | --- | --- | --- | --- | --- | --- |
|  |  |  | Number | Male | Female | Mean Age (SD) | Number | Male | Female | Mean Age (SD) |
| Zhang Y (2016)[23] | China | RCT | 31 | 17 | 14 | 55.0 ± 4.81 | 31 | 18 | 13 | 56.72 ± 3.57 |
| Xu L (2023)[24] | China | RCT | 30 | 18 | 12 | 64.29 ± 3.51 | 30 | 16 | 14 | 65.31 ± 3.62 |
| Tang Q (2022)[25] | China | RCT | 33 | 21 | 12 | 54.9 ± 13.1 | 34 | 20 | 14 | 56.5 ± 11.2 |
| Mu ZC (2021)[26] | China | RCT | 31 | 16 | 15 | 53.22 ± 1.78 | 31 | 17 | 14 | 52.48 ± 1.63 |
| Wang C (2022)[27] | China | RCT | 32 | 24 | 8 | 65.25 ± 9.21 | 31 | 24 | 7 | 60.65 ± 12.21 |
| Fu CX (2016)[28] | China | RCT | 30 | 19 | 11 | 59.7 ± 7.6 | 30 | 18 | 12 | 60.3 ± 8.4 |
| Tang LX (2018)[29] | China | RCT | 30 | 15 | 15 | 59.2 ± 8.5 | 30 | 14 | 16 | 59.5 ± 8.8 |
| Taylor-Piliae (2014)[44] | USA | RCT | 53 | 34 | 19 | 71.5 ± 10.3 | 48 | 23 | 25 | 68.2 ± 10.3 |
| Au-Yeung (2009)[15] | China | RCT | 59 | 33 | 26 | 61.7 ± 10.5 | 55 | 33 | 22 | 65.9 ± 10.7 |
| Zheng YN (2021)[14] | China | RCT | 30 | 24 | 6 | 63.50 ± 10.36 | 30 | 19 | 11 | 67.23 ± 9.15 |
| Rhayun Song (2021)[17] | Korea | RCT | 18 | 10 | 8 | 58.72 ± 17.13 | 16 | 11 | 5 | 57.18 ± 10.65 |
| Tian HL (2024)[16] | China | RCT | 25 | 21 | 4 | 58 ± 9 | 25 | 22 | 3 | 58 ± 12 |
| Taylor-Piliae (2012)[45] | USA | RCT | 16 | 10 | 6 | 72.8 ± 10.1 | 12 | 7 | 5 | 64.5 ± 10.9 |
| HoYoung Kim (2015)[46] | Korea | RCT | 11 | 7 | 4 | 53.45 ± 11.54 | 11 | 6 | 5 | 55.18 ± 10.20 |
| Jia WZ (2008)[30] | China | RCT | 18 | 11 | 7 | 47.70 ± 15.50 | 16 | 9 | 7 | 51.50 ± 16.37 |
| Chen YN (2022)[31] | China | RCT | 37 | 29 | 8 | 63.03 ± 7.21 | 36 | 26 | 10 | 63.30 ± 8.11 |
| Cong HZ (2022)[32] | China | RCT | 20 | 14 | 6 | 56.1 ± 7.59 | 20 | 12 | 8 | 53.6 ± 5.4 |
| Guo SL (2021)[33] | China | RCT | 14 | 6 | 8 | 64.71 ± 4.97 | 13 | 6 | 7 | 62.62 ± 5.32 |
| Sun YT (2021)[34] | China | RCT | 50 | 43 | 7 | 59.72 ± 9.59 | 38 | 29 | 9 | 59.45 ± 9.23 |
| Ling Y (2024)[35] | China | RCT | 21 | 17 | 4 | 54.14 ±<br>12.30 | 21 | 17 | 4 | 52.86 ±<br>14.84 |
| Zhou HY (2021)[36] | China | RCT | 50 | 29 | 21 | 53.32±<br>4.16 | 50 | 28 | 22 | 51.45±<br>4.09 |
| Lu LN (2023)[37] | China | RCT | 35 | 20 | 15 | 69.1±8<br>.5 | 35 | 22 | 13 | 69.5±8<br>.3 |
| Che P (2024)[38] | China | RCT | 43 | 28 | 15 | 63.10+<br>4.11 | 43 | 27 | 16 | 64.72+<br>3.65 |
| Xu J (2022)[39] | China | RCT | 28 | 17 | 11 | 62.04±<br>10.26 | 28 | 22 | 6 | 65.07±<br>9.82 |
| Qian C (2024)[40] | China | RCT | 36 | 19 | 17 | 60.48+<br>7.89 | 38 | 22 | 16 | 61.78+<br>8.06 |
| Xue XX (2024)[41] | China | RCT | 20 | 9 | 11 | 53.400<br>±5.985 | 20 | 8 | 12 | 52.750<br>±6.332 |
| Liu W (2022)[42] | China | RCT | 20 | 13 | 7 | 52.95±<br>8.38 | 20 | 13 | 7 | 57.40±<br>9.16 |
| Fan J (2020)[43] | China | RCT | 21 | 14 | 7 | 59.19±<br>4.69 | 22 | 15 | 7 | 57.73±<br>5.55 |

**Table 2.**
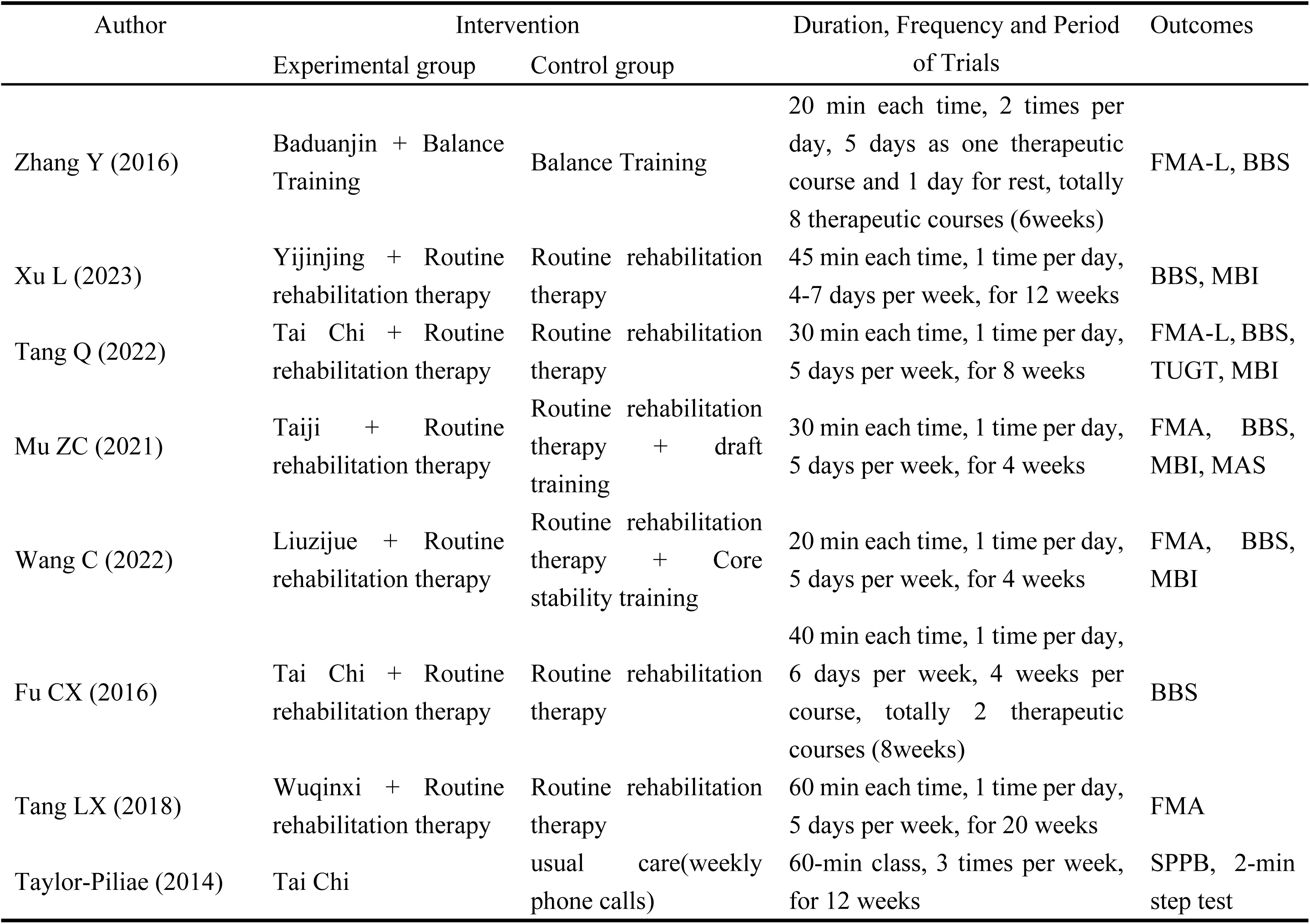
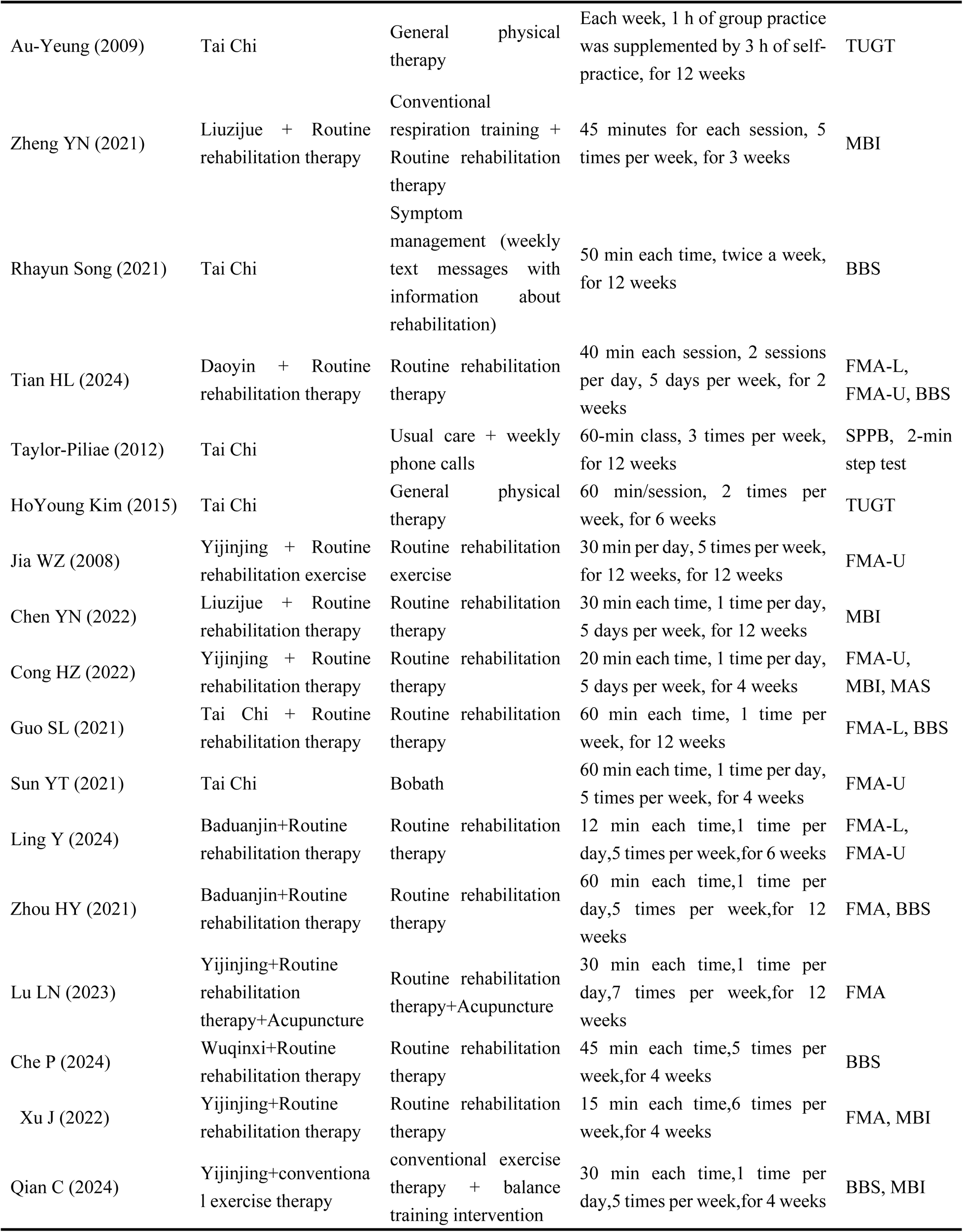
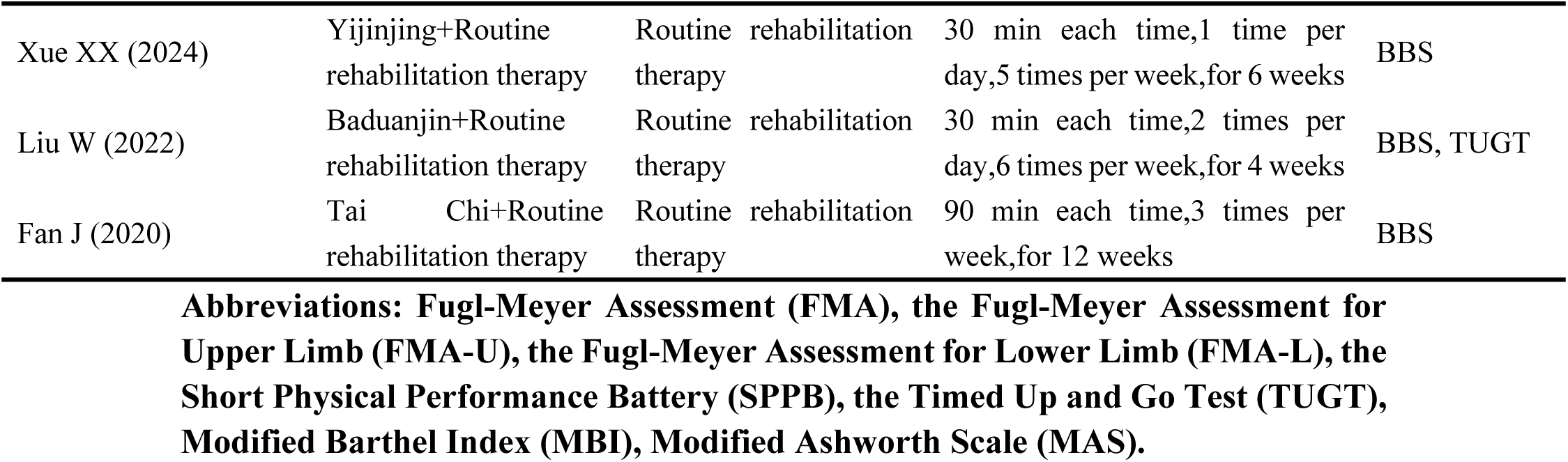
Forms of intervention included in studies.

| Author | Intervention |  | Duration, Frequency and Period of Trials | Outcomes |
| --- | --- | --- | --- | --- |
|  | Experimental group | Control group |  |  |
| Zhang Y (2016) | Baduanjin + Balance Training | Balance Training | 20 min each time, 2 times per day, 5 days as one therapeutic course and 1 day for rest, totally 8 therapeutic courses (6weeks) | FMA-L, BBS |
| Xu L (2023) | Yijinjing + Routine rehabilitation therapy | Routine rehabilitation therapy | 45 min each time, 1 time per day, 4-7 days per week, for 12 weeks | BBS, MBI |
| Tang Q (2022) | Tai Chi + Routine rehabilitation therapy | Routine rehabilitation therapy | 30 min each time, 1 time per day, 5 days per week, for 8 weeks | FMA-L, BBS, TUGT, MBI |
| Mu ZC (2021) | Taiji + Routine rehabilitation therapy | Routine rehabilitation therapy + draft training | 30 min each time, 1 time per day, 5 days per week, for 4 weeks | FMA, BBS, MBI, MAS |
| Wang C (2022) | Liuzijue + Routine rehabilitation therapy | Routine rehabilitation therapy + Core stability training | 20 min each time, 1 time per day, 5 days per week, for 4 weeks | FMA, BBS, MBI |
| Fu CX (2016) | Tai Chi + Routine rehabilitation therapy | Routine rehabilitation therapy | 40 min each time, 1 time per day, 6 days per week, 4 weeks per course, totally 2 therapeutic courses (8weeks) | BBS |
| Tang LX (2018) | Wuqinxi + Routine rehabilitation therapy | Routine rehabilitation therapy | 60 min each time, 1 time per day, 5 days per week, for 20 weeks | FMA |
| Taylor-Piliae (2014) | Tai Chi | usual care(weekly phone calls) | 60-min class, 3 times per week, for 12 weeks | SPPB, 2-min step test |
| Au-Yeung (2009) | Tai Chi | General physical therapy | Each week, 1 h of group practice was supplemented by 3 h of self-practice, for 12 weeks | TUGT |
| Zheng YN (2021) | Liuzijue + Routine rehabilitation therapy | Conventional respiration training + Routine rehabilitation therapy | 45 minutes for each session, 5 times per week, for 3 weeks | MBI |
| Rhayun Song (2021) | Tai Chi | Symptom management (weekly text messages with information about rehabilitation) | 50 min each time, twice a week, for 12 weeks | BBS |
| Tian HL (2024) | Daoyin + Routine rehabilitation therapy | Routine rehabilitation therapy | 40 min each session, 2 sessions per day, 5 days per week, for 2 weeks | FMA-L, FMA-U, BBS |
| Taylor-Piliae (2012) | Tai Chi | Usual care + weekly phone calls | 60-min class, 3 times per week, for 12 weeks | SPPB, 2-min step test |
| HoYoung Kim (2015) | Tai Chi | General physical therapy | 60 min/session, 2 times per week, for 6 weeks | TUGT |
| Jia WZ (2008) | Yijinjing + Routine rehabilitation exercise | Routine rehabilitation exercise | 30 min per day, 5 times per week, for 12 weeks, for 12 weeks | FMA-U |
| Chen YN (2022) | Liuzijue + Routine rehabilitation therapy | Routine rehabilitation therapy | 30 min each time, 1 time per day, 5 days per week, for 12 weeks | MBI |
| Cong HZ (2022) | Yijinjing + Routine rehabilitation therapy | Routine rehabilitation therapy | 20 min each time, 1 time per day, 5 days per week, for 4 weeks | FMA-U, MBI, MAS |
| Guo SL (2021) | Tai Chi + Routine rehabilitation therapy | Routine rehabilitation therapy | 60 min each time, 1 time per week, for 12 weeks | FMA-L, BBS |
| Sun YT (2021) | Tai Chi | Bobath | 60 min each time, 1 time per day, 5 times per week, for 4 weeks | FMA-U |
| Ling Y (2024) | Baduanjin+Routine rehabilitation therapy | Routine rehabilitation therapy | 12 min each time, 1 time per day, 5 times per week, for 6 weeks | FMA-L, FMA-U |
| Zhou HY (2021) | Baduanjin+Routine rehabilitation therapy | Routine rehabilitation therapy | 60 min each time, 1 time per day, 5 times per week, for 12 weeks | FMA, BBS |
| Lu LN (2023) | Yijinjing+Routine rehabilitation therapy+Acupuncture | Routine rehabilitation therapy+Acupuncture | 30 min each time, 1 time per day, 7 times per week, for 12 weeks | FMA |
| Che P (2024) | Wuqinxi+Routine rehabilitation therapy | Routine rehabilitation therapy | 45 min each time, 5 times per week, for 4 weeks | BBS |
| Xu J (2022) | Yijinjing+Routine rehabilitation therapy | Routine rehabilitation therapy | 15 min each time, 6 times per week, for 4 weeks | FMA, MBI |
| Qian C (2024) | Yijinjing+conventional exercise therapy | conventional exercise therapy + balance training intervention | 30 min each time, 1 time per day, 5 times per week, for 4 weeks | BBS, MBI |
| Xue XX (2024) | Yijinjing+Routine rehabilitation therapy | Routine rehabilitation therapy | 30 min each time,1 time per day,5 times per week,for 6 weeks | BBS |
| Liu W (2022) | Baduanjin+Routine rehabilitation therapy | Routine rehabilitation therapy | 30 min each time,2 times per day,6 times per week,for 4 weeks | BBS, TUGT |
| Fan J (2020) | Tai Chi+Routine rehabilitation therapy | Routine rehabilitation therapy | 90 min each time,3 times per week,for 12 weeks | BBS |

**Abbreviations: Fugl-Meyer Assessment (FMA), the Fugl-Meyer Assessment for Upper Limb (FMA-U), the Fugl-Meyer Assessment for Lower Limb (FMA-L), the Short Physical Performance Battery (SPPB), the Timed Up and Go Test (TUGT), Modified Barthel Index (MBI), Modified Ashworth Scale (MAS).**

### Quality assessment and risk of bias of included studies

**Figs 2 and 3** showed the quality assessment and risk of bias of literatures used for analysis. Baseline data for participants were reported in every study. There were 22 studies provided details on random sequence generation[14, 16, 17, 23–27, 29, 31–38, 40–43], and 9 studies implemented allocation concealment[14, 16, 26, 32, 34, 41, 42, 44, 45]. Informed consent was obtained in 28 studies. Eight studies reported participant dropouts[15, 25, 31–35, 38, 40–42, 44, 46], and only 3 studies conducted intention-to-treat analyses[15, 34, 44]. Blinding was applied to outcome assessments in 14 studies[14–17, 25, 27, 28, 30, 33, 35, 38, 40, 44, 45]. Ten studies used statistical methods to estimate sample sizes[17, 25–27, 31, 32, 34, 38, 40, 41]. Due to the inherent challenges of blinding participants and researchers in exercise therapy trials, and the minimal potential impact of missing blinding on outcomes. Therefore, all of them are included in the low-risk segment in this study. Based on the GRADE evaluation of the evidence, 18 studies were downgraded, and none of the studies encountered upgrade evaluation. The literature was downgraded due to the lack of an explicit randomization scheme and uncertainty regarding the blinding of the evaluator. During the assessment, there were 10 low-risk articles[14, 16, 17, 25, 27, 33, 35, 38, 40, 45], 16 medium-risk articles[15, 23, 24, 26, 28, 30–32, 34, 36, 37, 41–44], and 2 high-risk articles[39, 46]. The high-risk designation was attributed to the absence of a defined corporate random assignment scheme and the lack of blinding for the evaluator.

**Fig 2.**
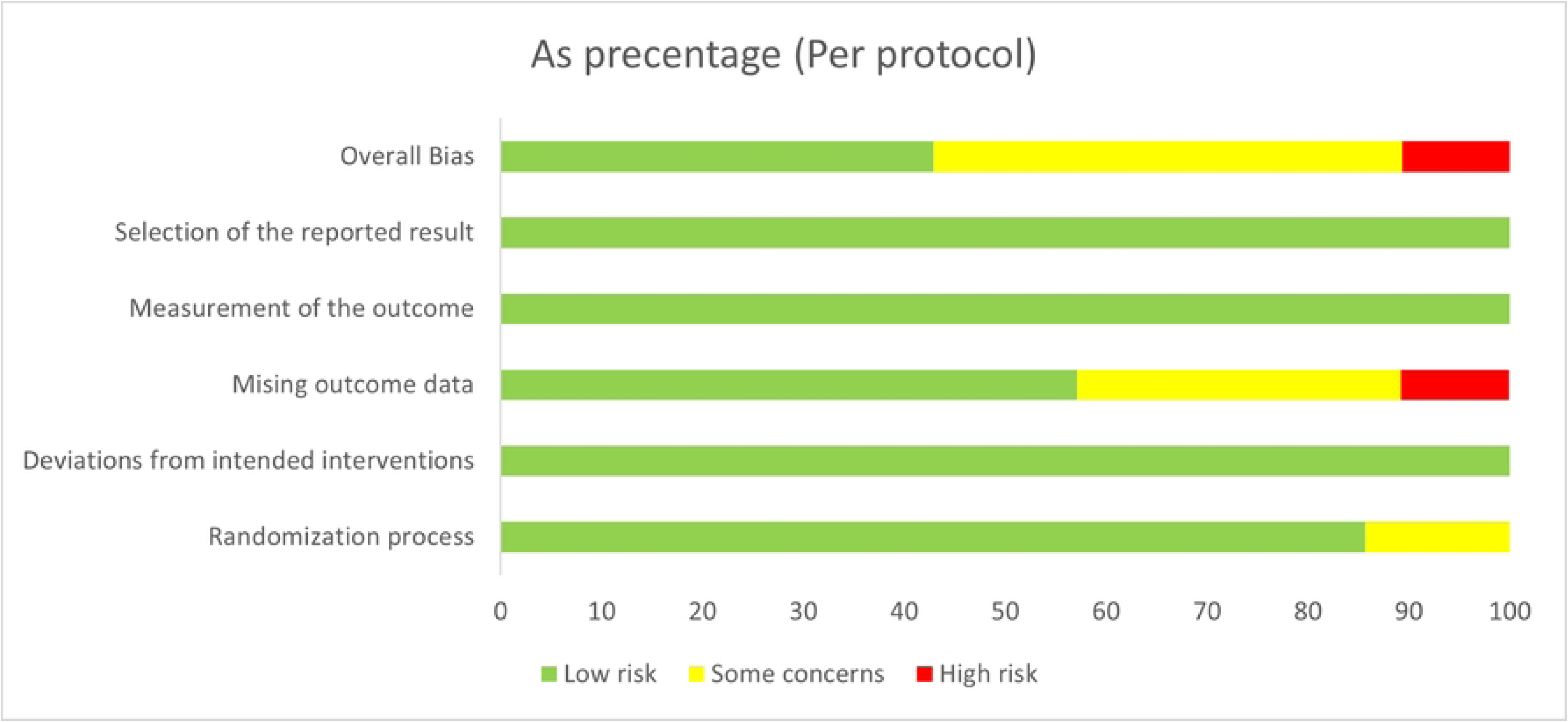
Risk of bias of the included studies.

**Fig 3.**
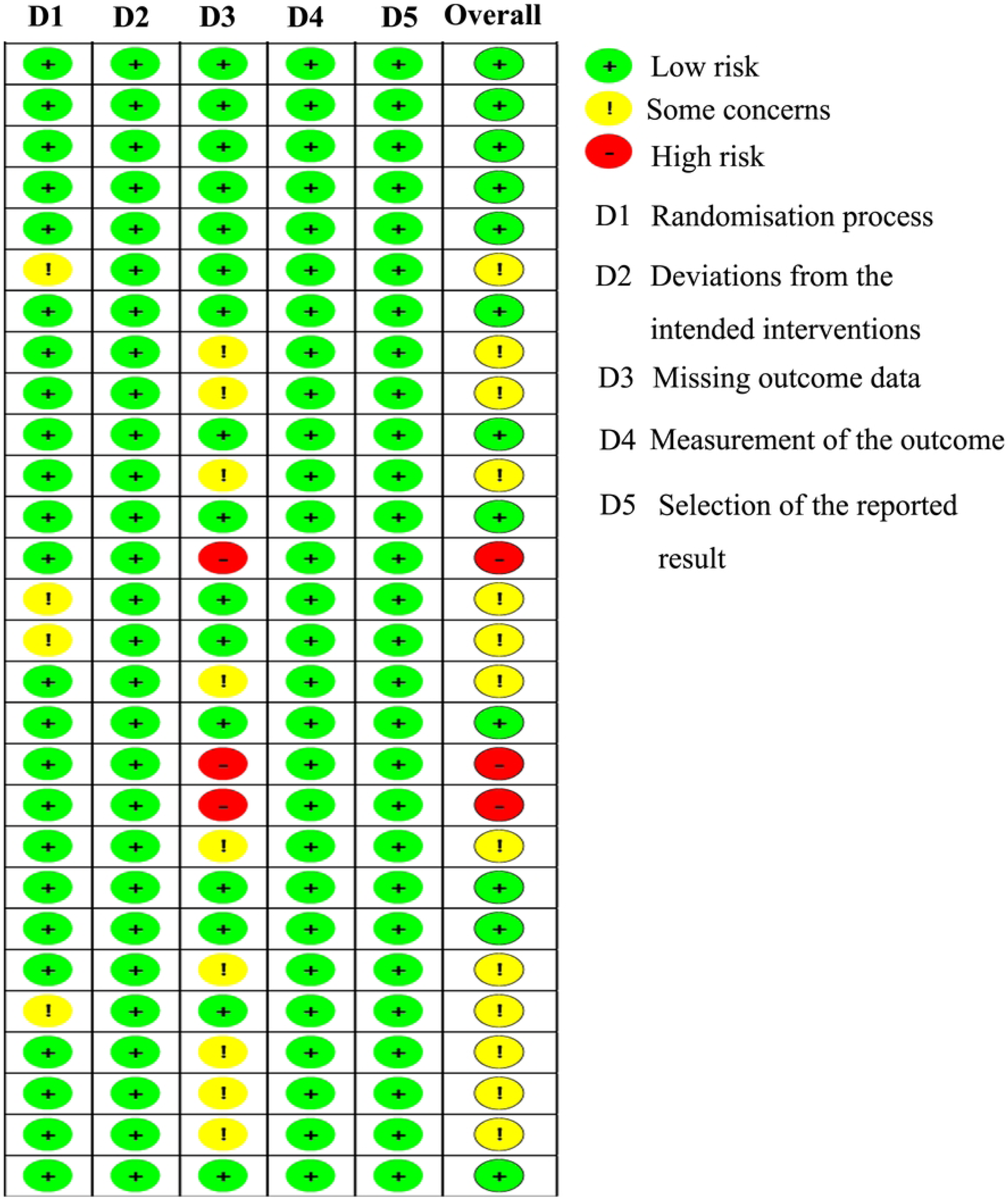
Risk of bias summary of the included studies.

### Effectiveness of TCEs for limb motor function rehabilitation after stroke

Fourteen studies utilized FMA(include FMA, FMA-L and FMA-U) to evaluate motor function in 874 stroke patients. Two of these studies included both FMA-L and FMA-U outcomes. **Table 3** presented detailed information. According to the meta-analysis, the experimental group had a markedly higher FMA score than the control group(SMD = 1.05, 95% CI: [0.70 to 1.04], *P* < 0.001, I^2^ = 85%). Given that various FMA subscales (FMA-L, FMA-U, and total FMA) were employed across the studies, a subgroup analysis was performed. The results indicated that the experimental group outperformed the control group in all FMA categories, including total FMA (MD = 9.96, 95% CI: [5.08 to 14.85], *P* < 0.001, I^2^ = 89%), FMA-L (MD = 3.74, 95% CI: [2.48 to 5.01], *P* < 0.001, I^2^ = 49%), and FMA-U (MD = 5.63, 95% CI: [3.91 to 7.34], *P* < 0.001, I^2^ = 35%). Additionally, we examined the impact of various types of TCEs. Significant improvements in FMA scores were observed for Tai Chi (SMD = 0.77, 95% CI: [0.51 to 1.04], *P* < 0.001, I^2^ = 42%), Yijinjing (SMD = 2.14, 95% CI: [1.81 to 2.48], *P* < 0.001, I^2^ = 44%), and Baduanjin (SMD = 0.86, 95% CI: [0.63 to 1.09], *P* < 0.001, I^2^ = 37%) compared to the control group, while Daoyin(SMD = 0.21, 95% CI: [-0.18 to 0.61], *P* = 0.29, I^2^ = 14%) did not show any notable differences. Analysis of the intervention duration indicated that both short-term (1-8 weeks, SMD = 0.72, 95% CI: [0.38 to 1.05], *P* < 0.001, I^2^ = 77%) and mid-term (8-16 weeks, SMD = 1.72, 95% CI: [0.96 to 2.48], *P* < 0.001, I^2^ = 86%) treatments resulted in significantly higher FMA scores for the experimental group, in comparison to the control group.

**Table 3.**
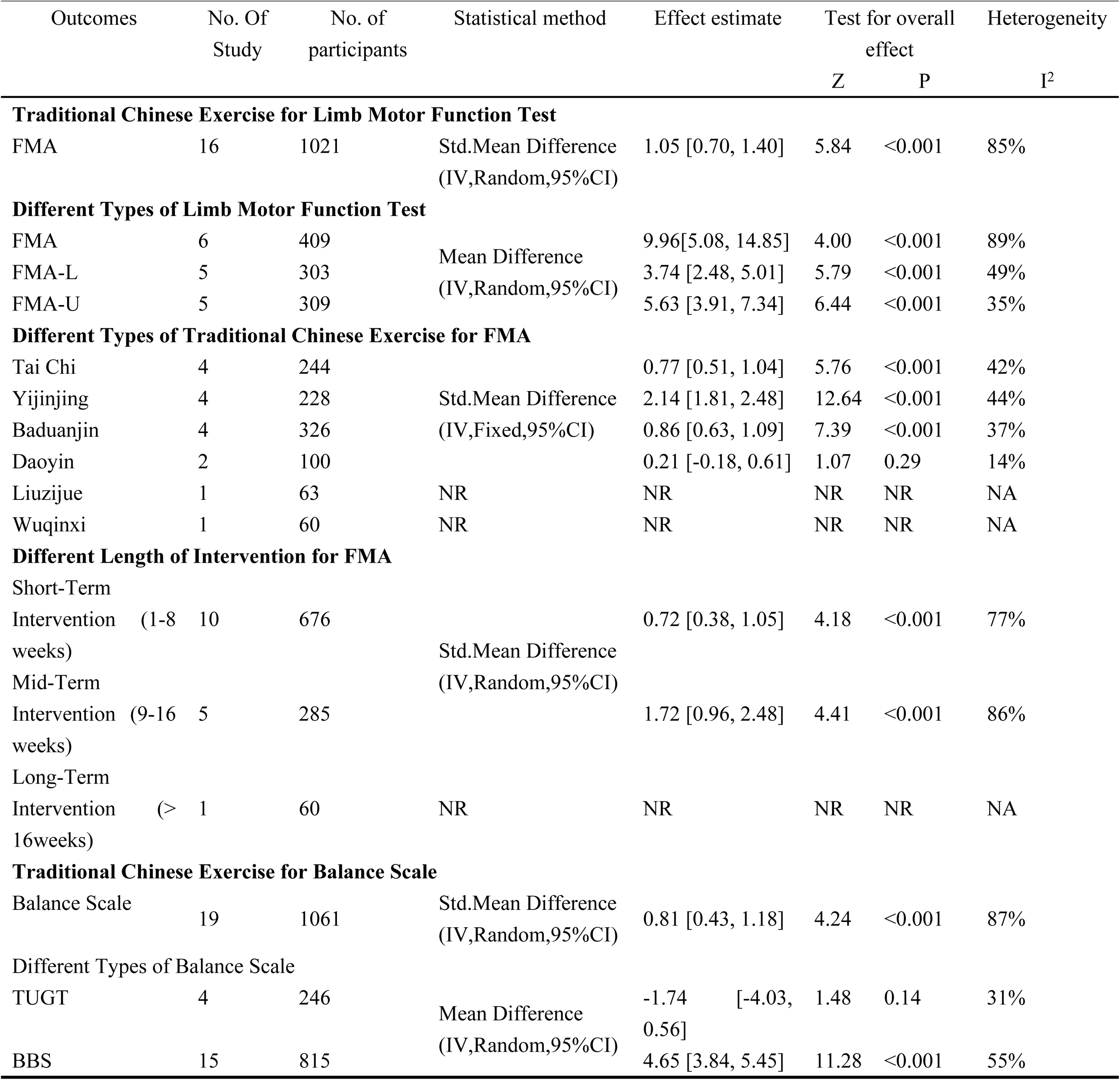
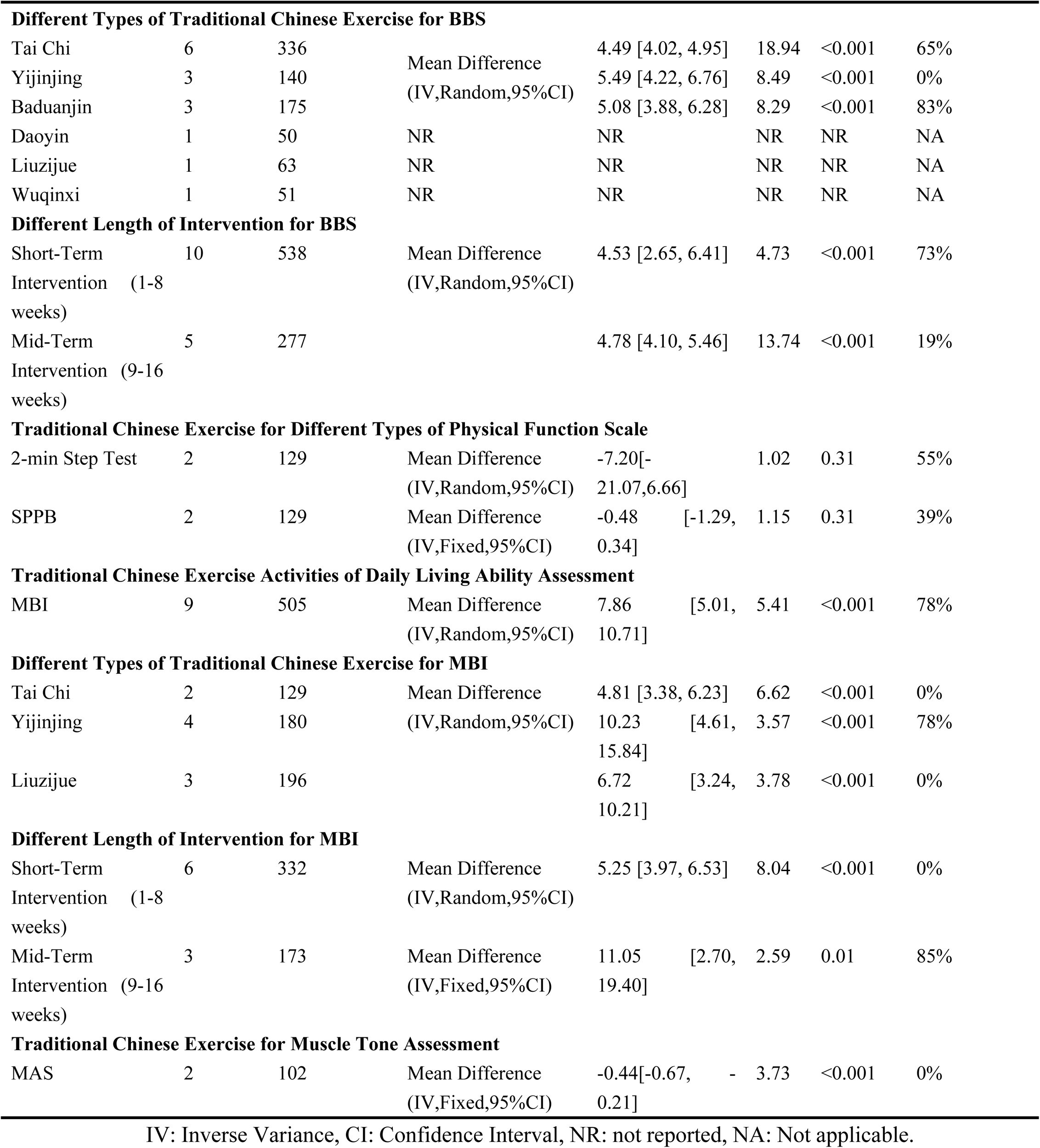
Outcome measures and results for included studies.

### Effectiveness of TCEs for different types of physical function scale

Physical function was assessed using the SPPB and 2-minute Step Test in two studies, involving a total of 129 participants. The meta-analysis revealed no notable differences in the SPPB (MD = -0.48, 95% CI: [-1.29 to 0.34], *P* = 0.25) or 2-minute Step Test (MD = -0.62, 95% CI: [-1.82 to 0.57], *P* = 0.31) scores between the experimental and control groups (**Table 3**).

### Effectiveness of TCEs for limb balance function rehabilitation after stroke

Seventeen studies assessed limb balance function among 940 stroke patients via TUGT and/or BBS (Table 3). The meta-analysis showed the experimental group had significantly higher balance scores than the control group (SMD = 0.81, 95% CI: [0.43 to 1.18], P < 0.001, I² = 87%). Given the use of two distinct balance scales, subgroup analysis was conducted. Results showed no significant difference in TUGT scores between the groups (MD = -1.74, 95% CI: [-4.03 to 0.56], *P* = 0.14), but the experimental group scored significantly higher on the BBS (MD = 4.65, 95% CI: [3.84 to 5.45], *P* < 0.001, I^2^ = 55%). Compared to the control group, Tai Chi(SMD = 4.49, 95% CI: [4.02 to 4.95], *P* < 0.001, I^2^ = 65%), Yijinjing (MD = 5.49, 95% CI: [4.22 to 6.76], *P* < 0.001, I^2^ = 0%) and Baduanjin (MD = 5.08, 95% CI: [3.88 to 6.28], *P* < 0.001, I^2^ = 83%) were observed to have significantly improved BBS scores, as revealed by the subgroup analysis based on TCE type. Moreover, subgroup analysis by intervention duration revealed that both short-term (1-8 weeks, MD = 4.53, 95% CI: [2.65 to 6.41], *P* < 0.001, I^2^ = 73%) and mid-term (8-16 weeks, MD = 4.78, 95% CI: [4.10 to 5.46], *P* < 0.001, I^2^ = 19%) interventions led to significant improvements in BBS scores in the experimental group, in contrast to the control group.

### Effectiveness of TCEs activities of daily living ability assessment

**Table 3** presents nine studies that used the MBI to evaluate daily living activities in 505 individuals who had strokes. The meta-analysis revealed that the experimental group had significantly higher MBI scores than the control group (MD = 7.28, 95% CI: [4.98 to 9.57], P < 0.001, I² = 64%). Subgroup analysis revealed that Tai Chi (MD = 4.81, 95% CI: [3.38 to 6.23], *P* < 0.001, I^2^ = 0%), Yijinjing (MD = 10.23, 95% CI: [4.61 to 15.84], *P* < 0.001, I^2^ = 78%), and Liu Zi Jue (MD = 6.72, 95% CI: [3.24 to 10.21], *P* < 0.001, I^2^ = 0%) were associated with significantly higher MBI scores compared to control interventions. Subgroup analysis based on intervention duration further demonstrated that MBI scores were significantly improved in the experimental group, both for short-term (1-8 weeks, MD = 5.25, 95% CI: [3.97 to 6.53], *P* < 0.001, I^2^ = 0%) and mid-term interventions (8-16 weeks, MD = 11.05, 95% CI: [2.70 to 19.40], *P* = 0.01, I^2^ = 85%), when compared to the control group.

### Effectiveness of TCEs for muscle tone assessment

Two studies assessed muscle tone in 102 stroke patients using the MAS (**Table 3**). The meta-analysis showed that MAS scores were significantly lower in the experimental group compared to the control group (MD = -0.44, 95% CI: [-0.67 to -0.21], *P* < 0.001).

### Sensitivity and publication bias analysis for studies

The results by subgroup analysis showed that the heterogeneity for “FMA-L and FMA-U for limb motor function Test”, “different types of traditional Chinese exercise for FMA”, “Yijinjing for BBS”, “Mid-Term Intervention for BBS”, “Tai Chi and Liuzijue for MBI”, “short-term intervention for MBI”, and “TCEs for muscle tone assessment” were significantly reduced (all *I²* < 50%). However, other results still showed significant heterogeneity (*P* < 0.05, *I²* > 50%). A sensitivity analysis was performed on the results exhibiting high heterogeneity to identify the sources of variation and evaluate the stability of the findings. The analysis indicated that removing certain studies reduced the I² value significantly, and the results remained stable. As shown in **Table 4**, the sensitivity analysis results are summarized.

**Table 4.** Results of sensitivity analysis.

| No. Of Study | Original effect estimate | *P* | *I*<sup>2</sup> | Method of analysis | No. Of Study | Effect estimate after sensitivity analysis | *P* | *I*<sup>2</sup> |
| --- | --- | --- | --- | --- | --- | --- | --- | --- |
| **Traditional Chinese Exercise for The Limb Motor Function Test** |  |  |  |  |  |  |  |  |
| 16 | 1.05 [0.70, 1.40] | <0.001 | 85% | Lu LN 2023, Cong HZ 2022, Jia WZ 2008, Tian HL 2024, Wang C 2022, and Xu J 2022 | 6 | 0.84 [0.64, 1.03] | <0.001 | 34% |
| **Traditional Chinese Exercise for The Limb Motor Function with FMA** |  |  |  |  |  |  |  |  |
| 6 | 9.96[5.08, 14.85] | <0.001 | 89% | Mu ZC 2021, Wang C 2022, and Xu J 2022 | 3 | 12.34[9.85,14.84] | <0.001 | 24% |
| **Short-Term Intervention for The Limb Motor Function with FMA** |  |  |  |  |  |  |  |  |
| 10 | 0.72 [0.38, 1.05] | <0.001 | 77% | Cong HZ 2022, Tian HL 2024, and Wang C 2022 | 3 | 0.79 [0.55, 1.03] | <0.001 | 43% |
| **Mid-Term Intervention for The Limb Motor Function with FMA** |  |  |  |  |  |  |  |  |
| 5 | 1.72 [0.96, 2.48] | <0.001 | 86% | Lu LN 2023, and Zhou HY 2021 | 2 | 1.74 [1.27, 2.22] | <0.001 | 23% |
| **Traditional Chinese Exercise for BBS** |  |  |  |  |  |  |  |  |
| 15 | 4.65 [3.84, 5.45] | <0.001 | 55% | Zhang Y 2016 | 1 | 4.39 [3.71, 5.07] | <0.001 | 35% |
| **Tai Chi for Balance with BBS** |  |  |  |  |  |  |  |  |
| 6 | 4.49 [4.02, 4.95] | <0.001 | 65% | Fu CX 2016 | 1 | 4.43 [3.97, 4.90] | <0.001 | 49% |
| **Baduanjin for Balance with BBS** |  |  |  |  |  |  |  |  |
| 3 | 5.08 [3.88, 6.28] | <0.001 | 83% | Zhang Y 2016 | 1 | 3.92 [2.54, 5.30] | <0.001 | 0% |
| **Short-Term Intervention for Balance with BBS** |  |  |  |  |  |  |  |  |
| 10 | 4.53 [2.65, 6.41] | <0.001 | 73% | Fu CX 2016, and Zhang Y 2016 | 2 | 3.52 [2.17, 4.88] | <0.001 | 38% |
| **Traditional Chinese Exercise for MBI** |  |  |  |  |  |  |  |  |
| 9 | 7.86 [5.01, 10.71] | <0.001 | 78% | Xu J 2022 | 1 | 5.53 [4.37, 6.69] | <0.001 | 0% |
| **Yijinjing for MBI** |  |  |  |  |  |  |  |  |
| 4 | 10.23[4.61,15.84] | <0.001 | 78% | Xu J 2022 | 1 | 7.08 [4.64, 9.53] | <0.001 | 0% |
| **Mid-Term Intervention for MBI** |  |  |  |  |  |  |  |  |
| 3 | 11.05[2.70,19.40] | <0.001 | 85% | Xu J 2022 | 1 | 6.82 [4.08, 9.57] | <0.001 | 0% |

Funnel plots were used to assess publication bias. Since some studies used multiple scales to assess the same outcome, they were counted more than once in the publication analysis. We assessed the publication bias for the Limb Motor Function Test (16 studies) and BBS (15 studies). The analysis revealed that both of them might have some publication bias (**Figs 4**, **Figs 5**).

**Fig 4.**
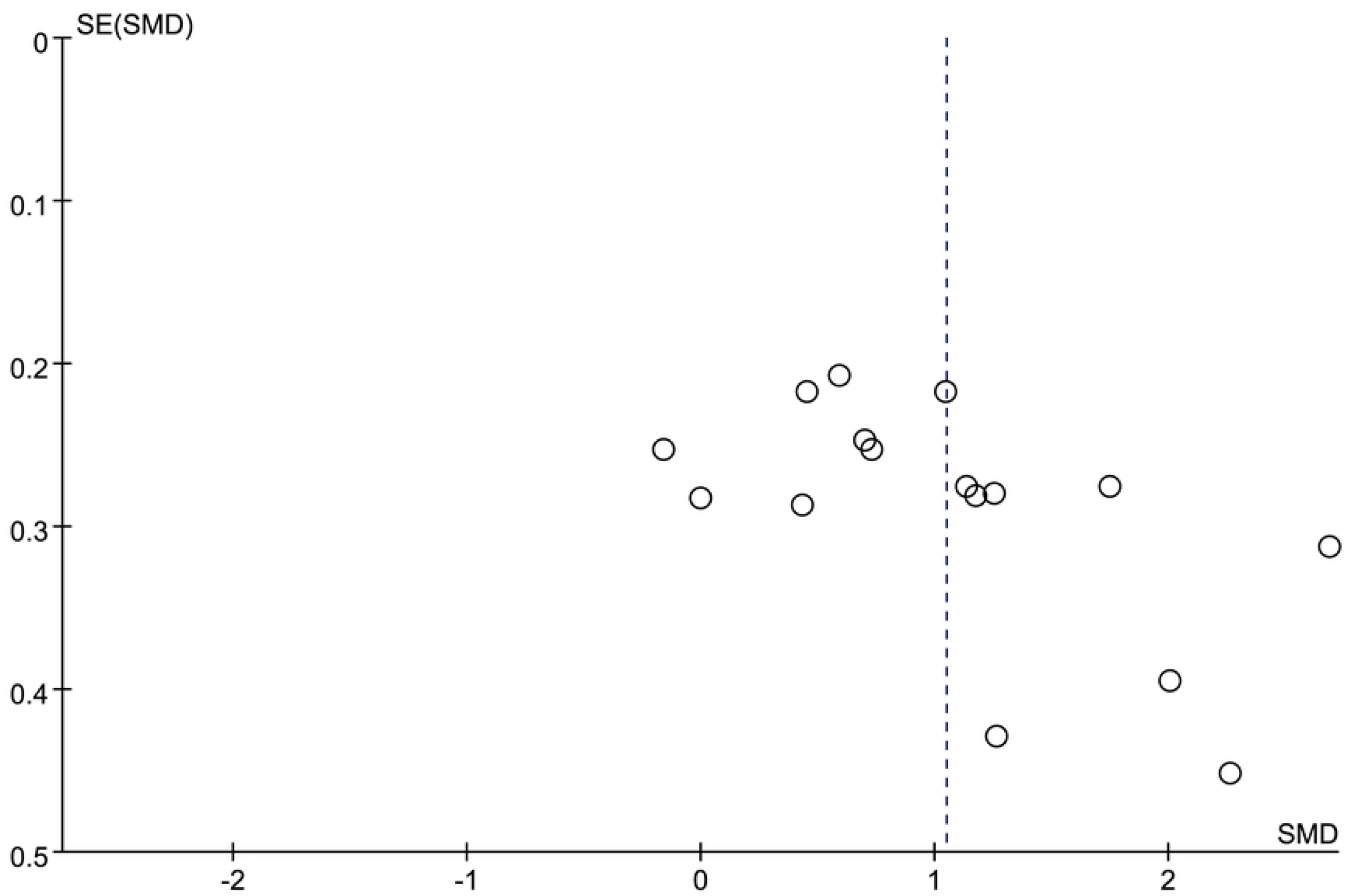
Funnel plot of the Fugl-Meyer Assessment scale regarding the impact of Traditional Chinese Exercises on post-stroke motor dysfunction.

**Fig 5.**
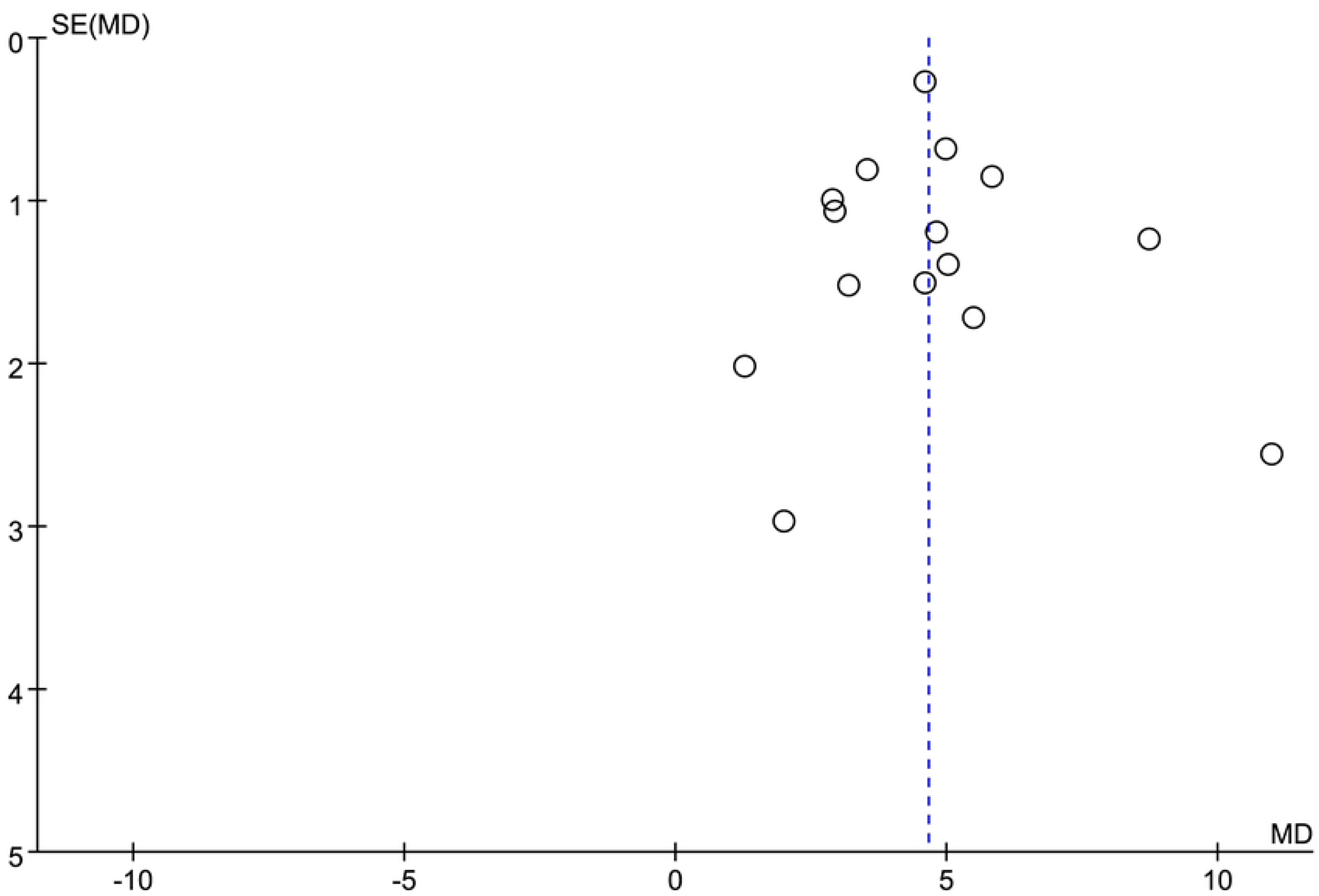
Funnel plot of the BBS scale examining the impact of Traditional Chinese Exercises on post-stroke limb balance dysfunction.

## Discussion

This systematic review intended to thoroughly assess the effectiveness of TCEs in rehabilitating limb function in stroke patients. Although many systematic reviews and meta-analyses had examined the impact of TCEs on stroke rehabilitation[18–20], they didn’t comprehensively cover all types of TCEs.A systematic review of 28 articles was conducted in this study to explore the role of TCEs in post-stroke limb dysfunction rehabilitation. It included six types of TCEs, such as Tai Chi, Baduanjin, Yijinjing, Daoyin, Liuzijue, and Wuqinxi. Overall analysis demonstrated that TCEs markedly improved motor function, balance function, daily living ability and muscle tone in stroke patients. Results highlighted the significant therapeutic value of TCEs for limb dysfunction and daily living limitations in stroke patients.

This study used FMA scores as the outcome measure for evaluating motor function. Although significant heterogeneity was observed, the overall analysis revealed that TCEs contributed to an improvement in FMA scores among stroke patients. Subgroup analyses were executed to identify the sources of heterogeneity, with a focus on discrepancies in FMA types, TCE categories, and intervention durations. The results showed that TCE consistently improved motor function scores regardless of the scale (FMA, FMA-L, FMA-U) used. These results align with those reported by Ge L et al[20]. Subgroup analysis by type of TCEs demonstrated that Tai Chi, Yijinjing, and Baduanjin significantly enhanced FMA scores in stroke patients, consistent with the findings from systematic reviews by Zheng XH et al.[47] and Wei LR et al[48]. Subgroup analysis revealed that different TCEs were potential sources of heterogeneity. This might be related to the exercise style, intensity, and movement characteristics of these TCEs. Tai Chi, featuring gentle, slowed circular movements and a blend of hardness and softness, was a renowned traditional Chinese exercise. Much research had shown that it could enhance balance and reduce falls[46, 49], particularly in older adults with postural and gait disorders and in stroke patients[13, 50]. Yijinjing focused on muscle and bone exercise as well as body flexibility. It stretched and strengthened muscles and bones through actions like stretching, twisting, and bending. Yijinjing had shown significant rehabilitation effects for joint functional disorders liked osteoarthritis and cervical spondylosis[51, 52], and it could also improve fatigue and sleep[53]. It was similarly valuable in the rehabilitation of stroke patients[11]. However, relevant English literature was still lacking, and more high-quality RCTs were needed for further investigation. Baduanjin enhanced balance and motor function in stroke patients[12, 54]. It centered on body stretching and relaxation, using simple movements to coordinate internal organs and limbs, improving joint mobility and unblocking meridians. However, its intensity was lower than Yijinjing. It showed a unique advantage in stroke patients who couldn’t undergo high - intensity training.

Yijinjing were more effective than Baduanjin and Tai Chi in improving motor function impairments. However, no similar effects were observed in “Daoyin,” which differs from a previous systematic review[20], suggesting that the therapeutic effect of Daoyin on post-stroke motor dysfunction remains controversial. Furthermore, because only one study each included Liuzijue and Wuqinxi, they could not be further analyzed. There was insufficient high-quality RCT evidence regarding the therapeutic effects of “Daoyin”, “Liuzijue”, and “Wuqinxi” on motor dysfunction after a stroke, highlighting the need for further randomized controlled trials. This meta-analysis performed a subgroup analysis to account for differences in intervention duration and different limb motor function scoring scales in the included studies. The results showed that the benefits of different TCEs for stroke patients were not influenced by different scales (FMA, FMA-L, FMA-U). The intervention duration of the included studies ranged from 2 to 20 weeks and had no impact on observed outcomes. However, the heterogeneity of subgroup results remained high. To investigate the sources of heterogeneity more thoroughly, a sensitivity analysis revealed a marked decrease in heterogeneity after excluding questionable studies, with the results remaining relatively stable. Heterogeneity was frequently attributed to the studies conducted by Cong et al., Tian HL et al., Xu J et al., and Wang C et al. This might be due to differences in the intervention methods. Tian HL et al. intervention method was Daoyin[16]. Cong HZ et al[32]., and Xu J et al[39]. used seated Yijinjing, Wang C et al. was Liuzijue[27].

The overall evaluation of TCEs for stroke patients’ balance function showed that these exercises significantly improved balance test scores, which aligns with recent systematic reviews[55], despite considerable heterogeneity. Subgroup analysis confirmed that TCEs enhanced BBS scores, but no significant differences were observed in TUGT scores. The analysis of different types of TCE showed that Taijiquan, Yijinjing, and Baduanjin could significantly improve BBS scores. This was similar to the results of two previous systematic reviews[20, 56], but this study did not evaluate Yijinjing and Taiji. ***T***ai Chi, which involved whole-body movement to adjust the center of gravity and improve muscle and joint coordination[57]. Both short-term (1-8 weeks) and mid-term (9-16 weeks) interventions also significantly improved BBS scores in the experimental group. However, there was still high heterogeneity in these results. Sensitivity analysis revealed that heterogeneity was notably reduced after removing studies with questionable data. Fu CX et al.[28] and Zhang Y et al[23]. were a key source of heterogeneity. Fu CX included patients with stage IV limb function recovery and Berg score 21-40 for balance dysfunction, while Zhang Y administered balance therapy in the control group. This might have caused a large difference in BBS scores. Stroke not only impairs patients’ ability to perform daily activities (ADLs) but also affects their capacity to return to work and impacts their mental health. Therefore, improving ADL function is a critical goal in stroke rehabilitation. The MBI, a widely used scale in rehabilitation medicine, is commonly employed to assess ADL abilities in post-stroke patients[58]. This study demonstrated that TCEs significantly improved MBI scores, enhancing ADL function in stroke patients, which aligned with previous findings[20]. However, subgroup analysis for different types of TCEs was not conducted in previous study. Our findings showed that Tai Chi, Yijinjing, and Liuzijue led to a significant increase in MBI scores, and heterogeneity was observed only in Yijinjing. These findings suggested that differences in TCE types might contribute to the observed heterogeneity. Additionally, both short-term and mid-term interventions improved MBI scores in stroke patients, though mid-term interventions showed higher heterogeneity. The results of sensitivity analysis showed that after removing the study of Xu J[39], there was no heterogeneity in all the results. The study included patients with severe stroke hemiplegia, making the study the only source of heterogeneity.

This study also conducted a systematic review and meta-analysis of the MAS and Physical Function Scale. TCEs were found to significantly reduce MAS scores, with no notable heterogeneity, while the 2-minute Step Test and SPPB showed no significant differences. This suggests that further research is needed to better understand the effectiveness of TCEs on the Physical Function Scale. Regarding safety, five studies reported no adverse events and one study reported dizziness, suggesting that TCEs was generally safe.

### Limitations

Despite implementing rigorous inclusion and exclusion criteria to ensure the quality of the literature, this study still had several limitations: (1) This analysis focused solely on articles published in Chinese and English, which could potentially lead to language bias. (2) The variability in study methodologies increased the potential for bias. While 22 studies described random sequence generation, only nine implemented allocation concealment. There were 13 studies reporting withdrawal of participants, but only 3 studies reporting intention-to-treat analyses. In addition, 14 studies blinded the evaluators, and only 10 reported specific sample size estimates. (3) The inclusion of multiple types of TCEs, varying control measures, and differences in intervention timing, frequency, duration, and target age groups likely contributed to the significant heterogeneity observed in the results. (4) Most of the studies included in this review originated from Chinese journals and databases, indicating that broadening the global reach of TCEs is crucial for advancing research in this field. (5) The funnel plot for the BBS indicated a substantial risk of publication bias, potentially arising from editorial choices, reviewer preferences, and the motivations of researchers.

In summary, we offer the following recommendations for clinical studies investigating the role of TCEs in post-stroke motor dysfunction: Rigorous RCT designs are essential, incorporating proper sample size estimation, random sequence generation, allocation concealment, blinding of assessors, and intention-to-treat analysis. (2) To maintain the quality of patient training, it is important to account for the varying severity of stroke, which may affect training outcomes. Therefore, establishing baseline motor function standards for participants is necessary. This is particularly important for home-based training, where effective follow-up measures must be in place to monitor progress and make timely adjustments to training plans. (3) When evaluating potential adverse events caused by TCEs, researchers must adhere strictly to the study protocol and ensure data accuracy to maintain the reliability and validity of the study.

## Conclusion

TCEs had therapeutic effects on abnormal limb motor function, balance function, muscle tone and poor ability of daily living in stroke patients, while maintaining a favorable safety profile. Subgroup analysis revealed that different types of TCEs affect motor and balance functions differently. Tai Chi, Yijinjing, and Baduanjin could improve the movement, balance function and daily living ability of stroke patients. The timing of the intervention did not affect these outcomes. However, the existing evidence did not provide conclusive support to confirm whether the intervention group exhibited a greater improvement in physical function compared to the control group. Due to the limitations in methodology and sample sizes in existing studies, future research should emphasize the need for large, high-quality, multi-center randomized controlled trials. Such studies would provide more robust evidence to guide clinical treatment policies.

## Data Availability

All relevant data are within the manuscript and its Supporting Information files.

## Conflict of Interest

All authors declared that the research was conducted in the absence of any commercial or financial relationships that could be construed as a potential conflict of interest.

## Author contributions

**Writing – review & editing**: Wang Guo

**Validation:** Hong-yu Li

**Writing – original draft:** Wang Guo

**Formal analysis:** Hong-yu Li

**Software:** Song-qi Li and Xuan-ting Chen

**Methodology:** Wang Guo and Zhi-hao Wang

**Data curation and Project administration:** Qiang Tang

## Acknowledgements

We sincerely appreciate the support and assistance from all those who contributed to our article.

## Notes

### Competing Interest Statement

The authors have declared no competing interest.

### Funding Statement

The author(s) received no specific funding for this work.

### Author Declarations

This study is a systematic review, so ethics are not applicable.

